# Multi-organ complement deposition in COVID-19 patients

**DOI:** 10.1101/2021.01.07.21249116

**Authors:** Paolo Macor, Paolo Durigutto, Alessandro Mangogna, Rossana Bussani, Stefano D’Errico, Martina Zanon, Nicola Pozzi, PierLuigi Meroni, Francesco Tedesco

## Abstract

**Background:** Increased levels of circulating complement activation products have been reported in COVID-19 patients, but only limited information is available on complement involvement at tissue level. The mechanisms and pathways of local complement activation remain unclear.

**Methods:** We performed immunofluorescence analyses of autopsy specimens of lungs, kidney and liver from nine COVID-19 patients who died of acute respiratory failure. Snap-frozen samples embedded in OCT were stained with antibodies against complement components and activation products, IgG and spike protein of SARS-CoV-2.

**Findings:** Lung deposits of C1q, C4, C3 and C5b-9 were localized in the capillaries of the interalveolar septa and on alveolar cells. IgG displayed a similar even distribution, suggesting classical pathway activation. The spike protein is a potential target of IgG, but its uneven distribution suggests that other viral and tissue molecules may be targeted by IgG. Factor B deposits were also seen in COVID-19 lungs and are consistent with activation of the alternative pathway, whereas MBL and MASP-2 were hardly detectable. Analysis of kidney and liver specimens mirrored findings observed in the lung. Complement deposits were seen on tubules and vessels of the kidney with only mild C5b-9 staining in glomeruli, and on hepatic artery and portal vein of the liver.

**Interpretation:** Complement deposits in different organs of deceased COVID-19 patients caused by activation of the classical and alternative pathways support the multi-organ nature of the disease.

**Funding:** Grants from the Italian Ministry of Health (COVID-2020-12371808) to PLM and National Institutes of Health HL150146 to NP are gratefully acknowledged.

## Introduction

The vast majority of individuals infected by the new coronavirus SARS-CoV-2 manifest mild to moderate disease and usually recover within a few weeks. However, some of them, for unknown reasons, experience a severe form of disease and requires intensive care treatment [1, 2]. The respiratory tract is considered the main target of SARS-CoV-2 that infects epithelial cells in the trachea and bronchi and pneumocytes in the lungs, causing pneumonia that in more severe cases progresses to acute respiratory distress syndrome [3]. Nonetheless, other organs may also be involved including heart, kidneys, liver [4] due to the wide distribution of the virus receptor ACE-2 [5, 6]. Consistent with the multi-organ nature of this complex disease, analysis of a large number of COVID-19 patients has revealed that, while two third of severe cases manifests acute respiratory distress syndrome, one third develops heart and kidney failure, as well as liver dysfunction [7].

Hyper-inflammation is a common feature in symptomatic COVID-19 infection and is characterized by infiltration of inflammatory cells in the lungs and other infected organs, particularly evident in severe forms of the disease. This process is the result of dysregulated response of the innate immune system and is sustained by pro-inflammatory cytokines released by macrophages and other cells at tissue sites [8]. However, analysis of severe cases have shown that the clinical severity of the disease is not always associated with increased levels of pro-inflammatory cytokines and other markers of inflammation, such as C-reactive protein (CRP) [9].

Complement (C) has emerged as a potential key contributor to the development of inflammation and tissue damage in COVID-19 patients with the release of the pro-inflammatory peptides C3a and C5a that help to recruit leukocytes to the lung and other infected tissues and the assembly of the terminal complex that damage vascular endothelium and promotes thrombus formation [10, 11]. We have reported increased levels of C5a and sC5b-9 related to the severity of disease and not always associated with a parallel increase in acute phase proteins in COVID-19 patients group [12, 13]. Similar findings have been reported by Gao et al. in the preprint server MedRxiv [14] and, more recently, by Carvelli and colleagues who suggest the involvement of C5a-C5aR1 axis in the pathogenesis of SARS-CoV-2 infection and the potential benefit of the therapeutic blockade of this axis [15]. More direct evidence for the contribution of C to tissue damage was obtained from postmortem analysis of 2 lung and 3 skin biopsy specimens of COVID-19 patients that revealed deposits of C activation products C4d, C3d and C5b-9 in the lung inter-alveolar septal microvessels and in the skin vasculature [16]. The finding of MASP2 localized in the pulmonary inter-alveolar septa of one COVID-19 patient led Magro and coworkers to conclude that C is activated through the lectin pathway.

The aim of the present investigation is to analyze the distribution of early and late C components in the lung, kidney and liver of COVID-19 patients with the intent to clarify the pathway/s of C activation and to document the involvement of tissues other than lung as targets of C attack.

## Material and Methods

### Study group

The study group comprised 9 patients, 5 females and 4 males, aged 72 to 97 referred to the University Hospital in Trieste (Italy). Two patients were admitted to the Intensive Care Unit where they received intubation and mechanical ventilation. The remaining patients were followed in other medical sections of the Hospital including Infectious Diseases and Geriatrics wards or in nursing homes from which they were transferred to one of the Hospital wards when the clinical conditions deteriorated. The diagnosis of SARS-CoV-2 infection suspected on the basis of the clinical symptoms were confirmed by RT-PCR analysis of nasopharyngeal swab. This work was approved by the Ethical Committee of the Regione Friuli Venezia Giulia, Italy (Prot. N. 0025523 / P / GEN/ ARCS).

### Tissue sample collection

Limited autopsies from 9 SARS-CoV-2-positive and 3 SARS-CoV-2-negative cases with a similar age range were performed by an experienced pathologist and samples were collected from lungs, kidney and liver available for this study. Three or more tissue blocks were obtained from selected areas of the three organs and fixed for 24 hours in 10% buffered formalin. Part of these samples were paraffin-embedded and 3-µm thick sections were stained with haematoxylin and eosin for histological examination. Detailed analysis of tissue abnormalities in 41 autopsies including the nine autopsies reported in this work have been previously published [17].

### Immunofluorescence analysis

Part of the formalin-fixed tissue samples were snap-frozen and embedded in OCT medium (Diagnostic Division, Miles Inc, Tarrytown, NY, USA). Tissue sections of seven µm were stained with the following primary antibodies (5μg/ml): goat anti-human IgG (Sigma-Aldrich, Milan, Italy), goat anti-C1q and C4 (The Binding Site, Birmingham, UK), goat anti-C3 (Quidel, San Diego, CA, USA), and anti-Factor B (Cytotech, San Diego, CA, USA); rabbit anti-MBL (Sigma-Aldrich) and anti-SARS-CoV-2 Spike S2 (Sino Biological, Beijing, China); murine monoclonal antibody against C9 neo-antigen (aE11, kindly provided by prof. T.E. Mollnes, Oslo, Norway). The following FITC-conjugated secondary antibodies were used to reveal bound antibodies: rabbit anti-goat IgG (Sigma-Aldrich), goat anti-rabbit IgG and anti-mouse IgG (Dako, Jena, Germany). The slides were mounted with the Mowiol-based antifading medium (Sigma-Aldrich) and the Images were acquired with fluorescence microscope Leica DM2000 equipped with Leica DFC420 camera [18]. At least three random sections of tissue samples collected from each case were examined independently by three observers. Cases with moderate to strong staining intensity were considered as positive, while those with weak staining intensity were defined as weakly positive and all the others with undetectable staining were scored as negative.

## Results

### Clinical data

The study group included 9 patients, 5 females and 4 males, aged 72 to 97 **(Table 1)**. They all manifested symptoms related to pneumonia, which was confirmed by High Resolution Computer Tomography analysis in 8 of them. Radiological assessment could not be performed in one patient due to the very old age. Cancer and diabetes were the most frequent co-morbidities and 5 experienced also heart disease while 4 had neurological problems. Hypoxemia evaluated by blood gas analysis was particularly severe in all patients with PaO2 /FiO2 ratio of 100 or less. Increased levels of the inflammatory and coagulation markers CRP, ferritin and D-dimer and lymphopenia were commonly observed **(Table 2)**. As per hospital guidelines, the patients were treated with low molecular weight heparin and steroids to prevent clot formation and to control hyper-inflammation. Death occurred after 22±12 days of hospitalization (mean ± Standard Deviation) because of acute respiratory failure.

**Table 1.**
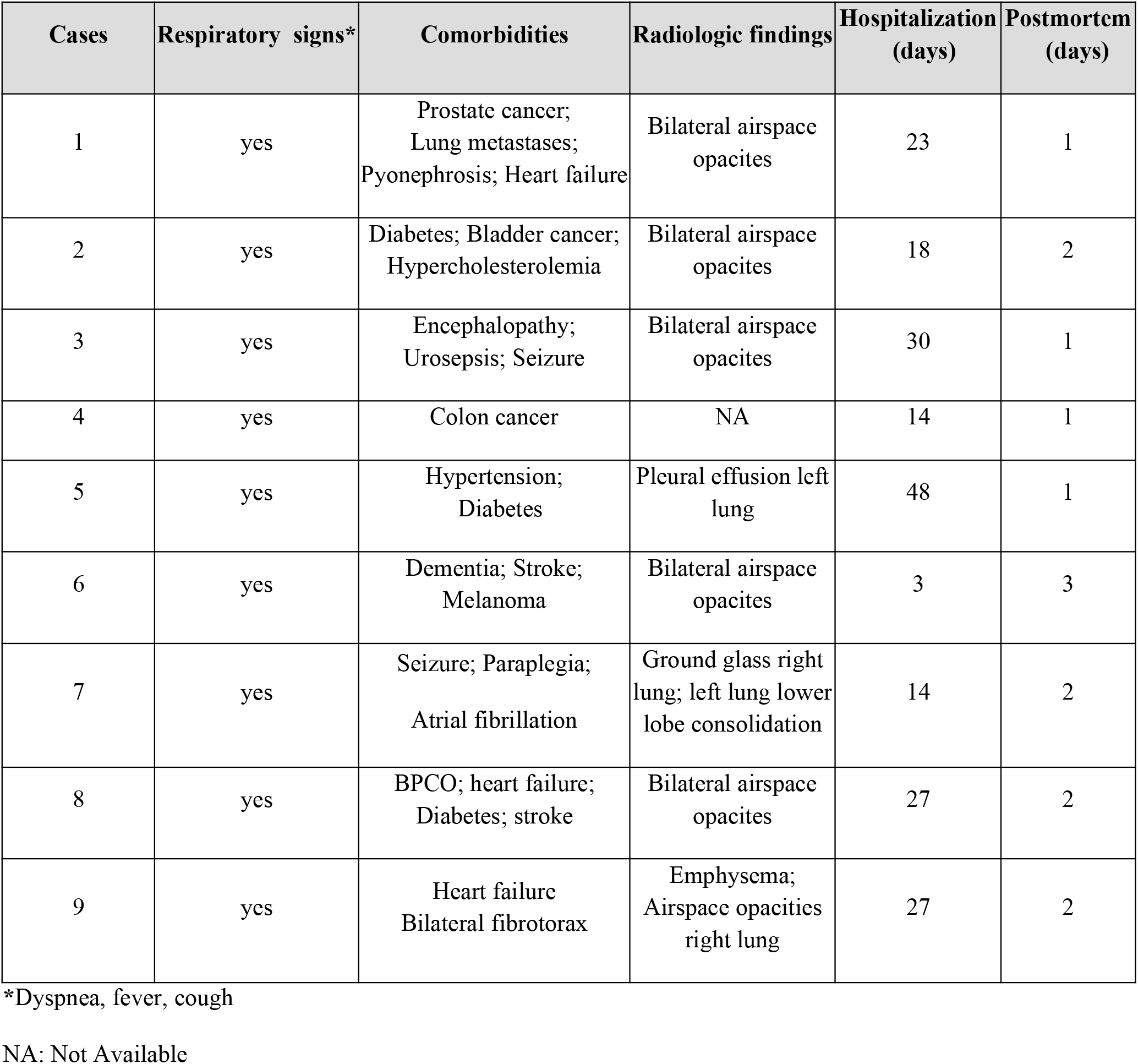
Clinical findings.

**Table 2.** Laboratory findings.

| Cases | D-dimer<br>(µg/L) | CRP<br>(mg/dL) | Ferritin<br>(µg/L) | White cells<br>(n x 10<sup>3</sup>/µL) | Neutrophils<br>(n x 10<sup>3</sup>/µL) | Lymphocytes<br>(n x 10<sup>3</sup>/µL) | Platelets<br>(n x 10<sup>3</sup>/µL) |
| --- | --- | --- | --- | --- | --- | --- | --- |
| 1 | 17180 | 15.6 | 2059 | 14.59 | 6.72 | 0.52 | 173 |
| 2 | 3020 | 30.6 | 2892 | 15.13 | 11.25 | 1.65 | 256 |
| 3 | 1330 | 30.2 | 90.9 | 12.98 | 9.59 | 2.45 | 2.84 |
| 4 | 1330 | 302,8 | 352 | 20.83 | 6.13 | 0.82 | 239 |
| 5 | NA | 9.1 | 85 | 12.68 | 11.21 | 0.43 | 461 |
| 6 | 6800 | 16.08 | 1608 | 5.57 | 3.89 | 0.76 | 231 |
| 7 | 2120 | 3.82 | 882 | 14.03 | 6.57 | 1.15 | 259 |
| 8 | 1800 | 210 | 755 | 14.19 | 13.51 | 0.31 | 83 |
| 9 | 430 | 15.7 | 1035 | 23.03 | 3.52 | 1.82 | 326 |
| Controls | <500 | <0.05 | 30-400 | 4.8-10.8 | 1.5-6.5 | 1.2-3.4 | 130-430 |

NA: Not Available

### C deposition in the lungs

Data from previous immunohistochemical studies have suggested the involvement of the lectin pathway in C activation in COVID-19 patients [14, 16]. To confirm these findings, we stained lung tissues with antibodies against MBL and, to our great surprise, we failed to detect tissue deposits of this protein, except for occasional faint staining of MBL in vascular thrombi. To determine if C was activated by other initiators of the lectin pathway such as ficolins and collectins, we examined the autopsy specimens for the presence of MASP-2 which is strictly required for the activation of the lectin pathway, but, as for MBL, the staining was negative (data not shown) We next investigated the activation of the classical pathway. Extensive deposition of C1q was found in the capillaries of the interalveolar septa and, to a lesser extent, on alveolar cells **(Figure 1)**. Since the distribution of C1q deposition was similar to that of IgG, C4 and C3 in all cases **(Figures 1)**, we concluded that the classical pathway is a common route of C activation in the lungs of our COVID-19 patients. We also looked into the activation of the alternative pathway by staining the sections for factor B, which was easily detectable (**Figure 1)**, and concluded that, in addition to the classical pathway, the alternative pathway must be playing an important role in C activation in these patients. In agreement with previous data by Magro et al., and Gao et al [14, 16], we found that the terminal complex C5b-9 localizes along the alveolar wall and on the interalveolar microvascular vessels **(Figures 1)**. The spike protein was detected on alveolar cells and interalveolar septal capillaries in 5 out of 9 specimens **(Figures 1)**. None of the control autopsy specimens showed deposits of C components and activation products, IgG and spike protein. To summarize, activation of both classical and alternative pathways documented by deposition of C1q, C4, C3 and factor B was observed in 6 cases, while activation of either pathway was found in two cases. C components and C activation products were undetectable in a single case, except for a mild staining for factor B and IgG **(Table 3)**.

**Table 3.** Complement deposition in lungs.

|  | **P1** | **P2** | **P3** | **P4** | **P5** | **P6** | **P7** | **P8** | **P9** |
| --- | --- | --- | --- | --- | --- | --- | --- | --- | --- |
| **IgG** | P | W | P | W | P | P | P | P | W |
| **C1q** | P | W | P | W | W | P | W | N | N |
| **MBL** | N | W | N | N | N | N | N | N | N |
| **Factor B** | P | P | P | P | P | P | N | W | W |
| **C4** | P | W | P | P | W | W | W | N | N |
| **C3** | P | W | P | P | P | P | W | W | N |
| **C5b-9** | W | W | W | W | P | N | N | N | N |

**P** Positive **W** Weakly positive **N** Negative

**Figure 1.**
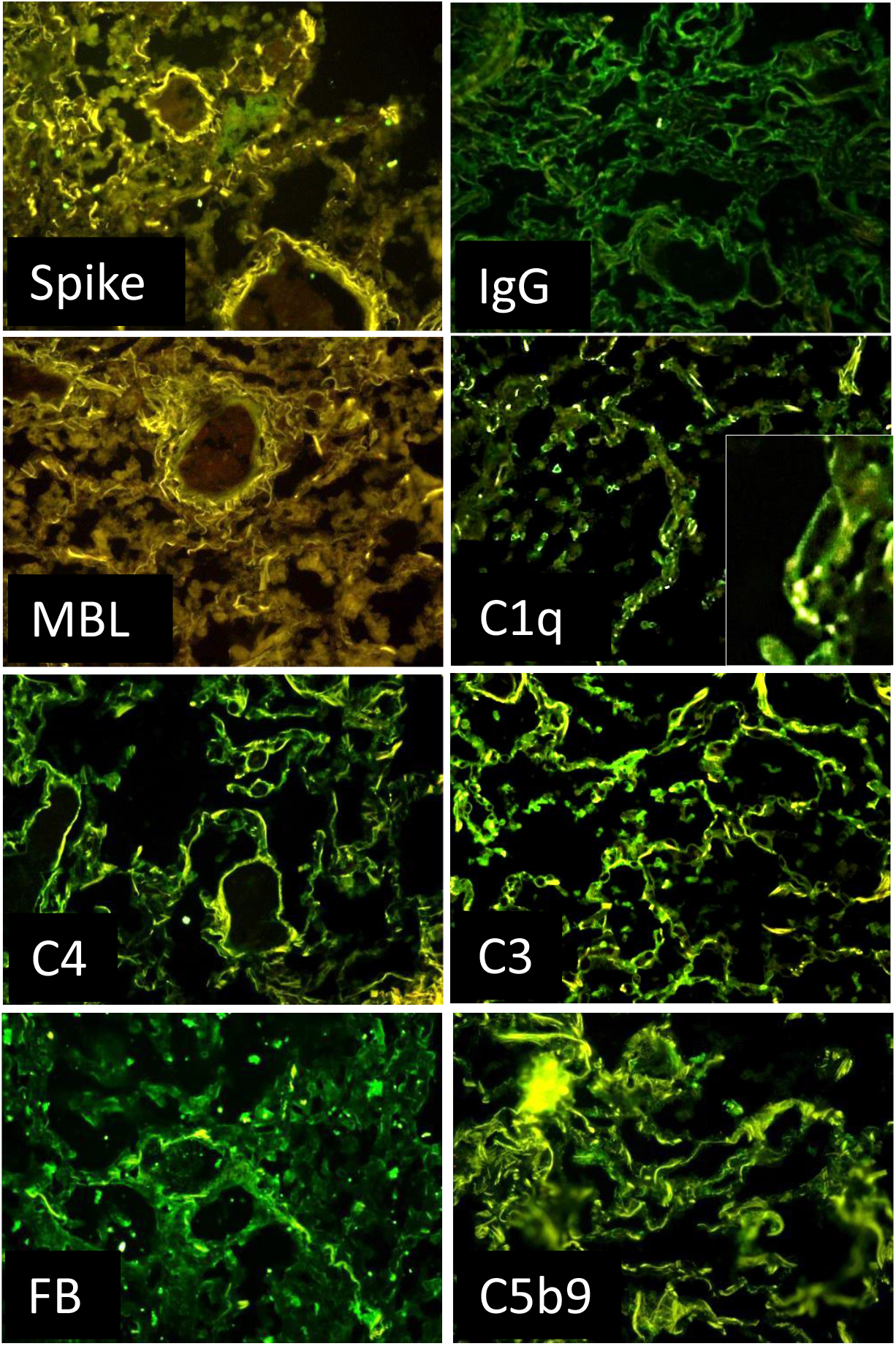
Detection of complement components in lung sections by immunofluorescence analysis. The panel shows representative immunofluorescence images obtained from the analysis of several sections of lung autopsy samples from 9 SARS-CoV-2-positive cases. The sections were stained to reveal the presence of spike protein, IgG, MBL, C1q, C4, C3, factor B (FB) and C5b-9 as explained in materials and methods and examined by 3 independent observers. Original magnification 200x.

### C deposition in kidneys and liver

Beyond the lungs, signs of C activation were searched in kidneys and liver, two organs that may be affected by the virus. The data summarized in **Table 4 and 5** show that C and IgG deposition patterns observed in kidneys **(Figure 2)**, liver **(Figure 3)**, were remarkably similar to those seen in the lung, suggesting that C activation may follow similar pathways in all these organs. The detection of C5b-9 both in kidney and liver supports the progression of C activation to completion in the majority of cases. A detailed analysis of kidney sections revealed the presence of IgG and C components on tubules, vessels and in the periglomerular areas and mild C5b-9 staining in glomeruli **(Figure 2)**. Staining for the spike protein was found in 5 out of 9 cases and was seen in the tubules and some small vessels, but was undetectable in the glomeruli **(Figure 2)**.

**Table 4.** Complement deposition in kidneys.

|  | P1 | P2 | P3 | P4 | P5 | P6 | P7 | P8 | P9 |
| --- | --- | --- | --- | --- | --- | --- | --- | --- | --- |
| **IgG** | P | W | W | W | P | P | W | P | W |
| **C1q** | P | W | P | W | W | P | N | N | N |
| **C4** | P | P | W | P | P | P | N | W | N |
| **C3** | P | P | P | P | P | P | W | W | W |
| **Factor B** | W | W | W | W | N | N | N | N | W |
| **C5b-9** | P | P | W | W | P | P | N | N | W |

P Positive W Weakly positive N Negative

**Table 5.**
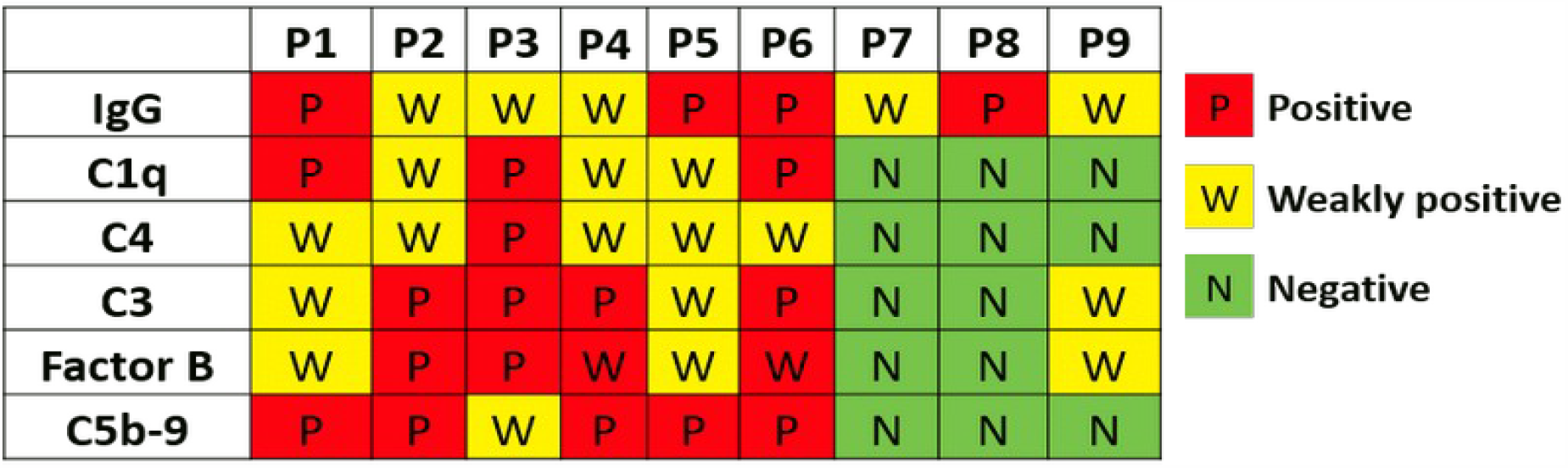
Complement deposition in livers.

**Figure 2.**
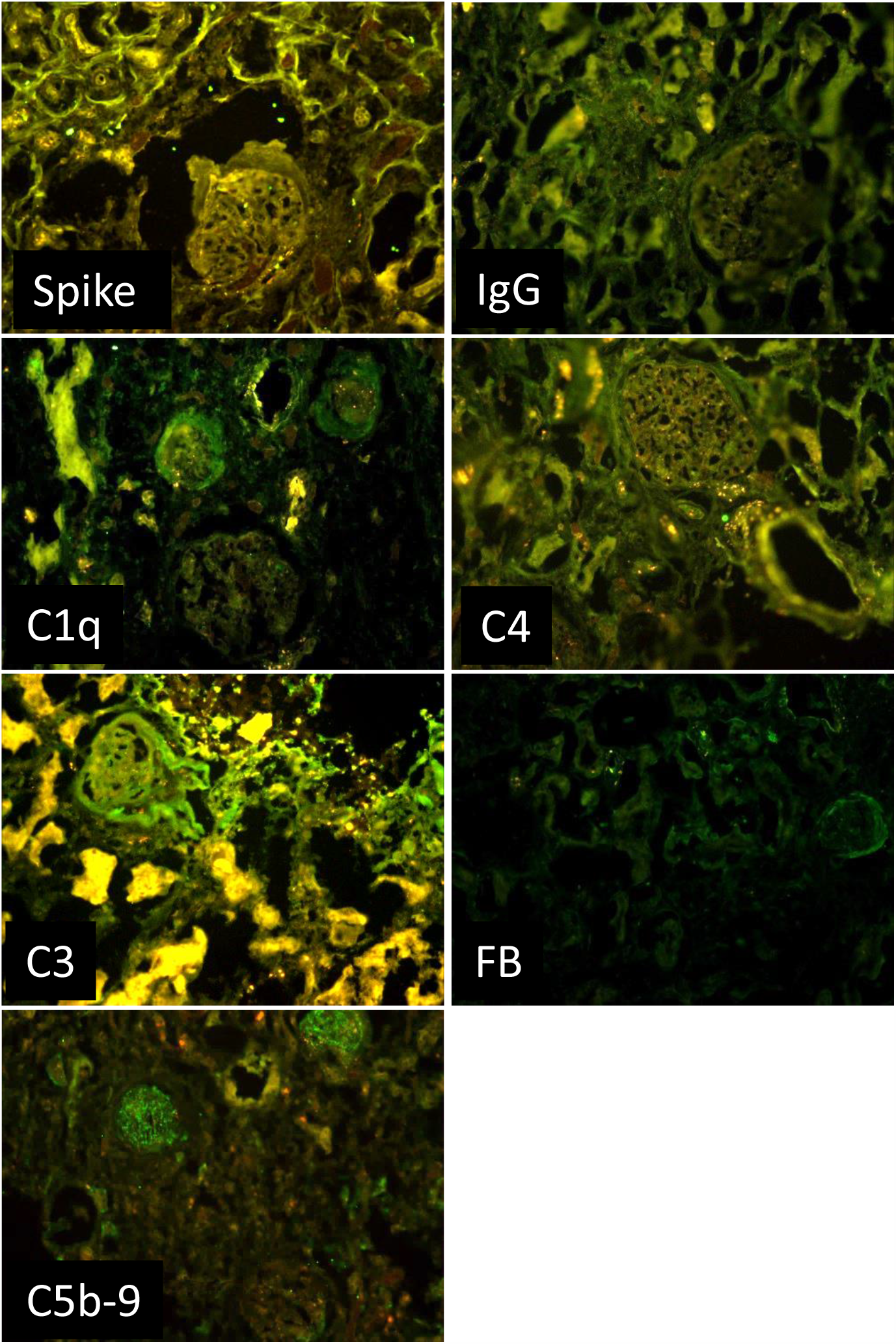
Detection of complement components in kidney sections by immunofluorescence analysis. The panel shows representative immunofluorescence images obtained from the analysis of several sections of kidney autopsy samples from 9 SARS-CoV-2-positive cases. See legend to Figure 1 for further details. Original magnification 200x.

**Figure 3.**
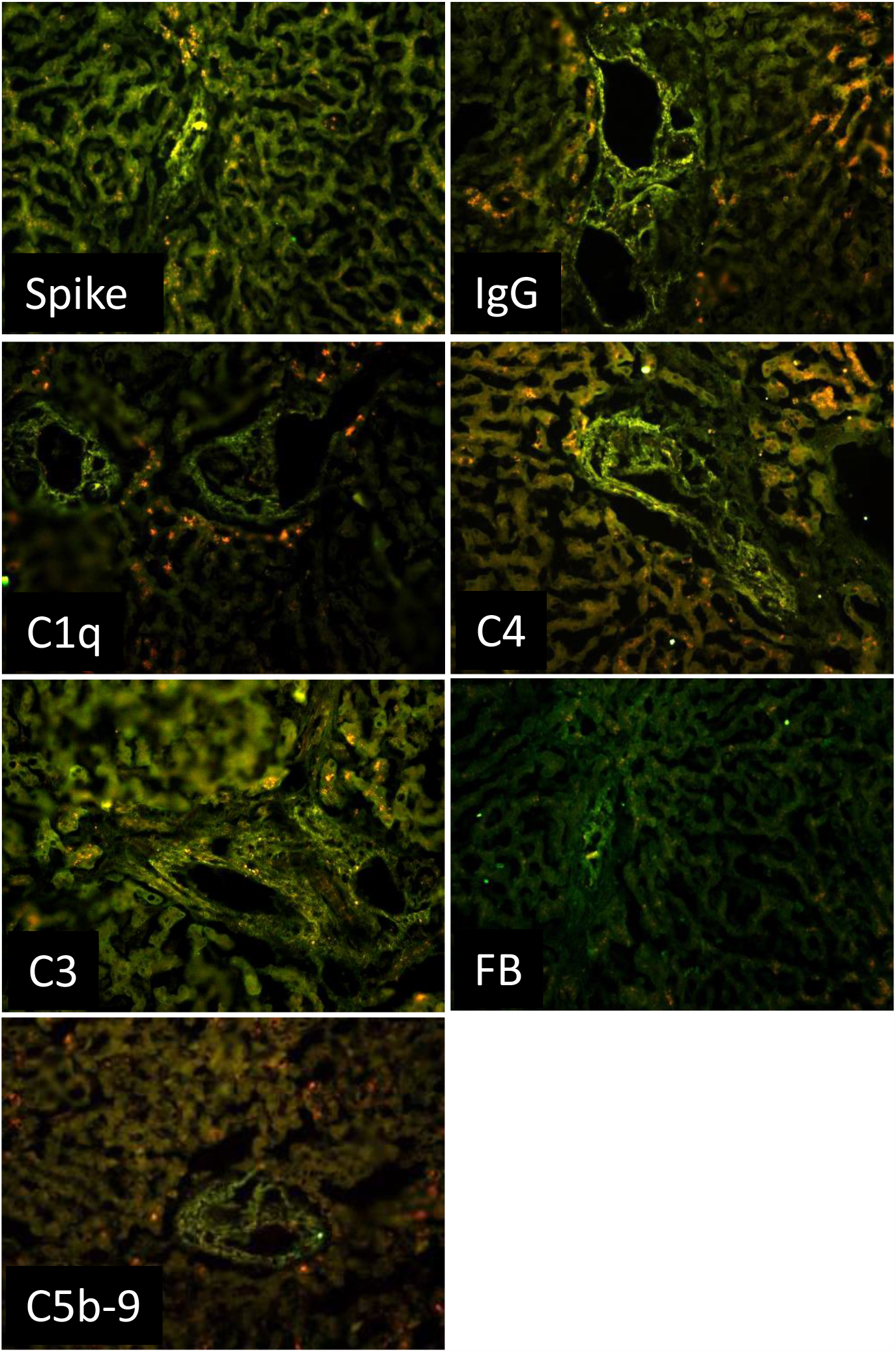
Detection of complement components in liver sections by immunofluorescence analysis. The panel shows representative immunofluorescence images obtained from the analysis of several sections of liver autopsy samples from 9 SARS-CoV-2-positive cases. See legend to Figure 1 for further details. Original magnification 200x.

As for the liver, the portal area represents the main site of C localization, which was observed primarily on the hepatic artery and portal vein **(Figure 2)**, supporting the vascular involvement in C-mediated tissue damage in COVID-19 patients. The spike protein was found to localize in the portal area of 5 out 9 cases.

## Discussion

C activation products detected in plasma of COVID-19 patients have recently emerged as useful markers to monitor the progression of the disease from early to advanced stages. However, it is important to acknowledge that these methods fail to report on the extent of local C activation, whose information is key to a better understanding of the mechanisms underlying tissue damage and disease progression. This work investigates C deposition in nine COVID-19 patients at the tissue level. Our results show that C deposits are present in the lungs, kidneys and liver, providing additional evidence for multi-organ injuries in the disease [4, 7, 19], and suggesting that comp-mediated damage can potentially affect other organs besides the lungs.

C deposits were found in the lungs of the majority of our patients, localized on the wall of the interalveolar septal capillaries and, to some extent, also on alveolar epithelial cells. Accumulation of C deposits in these anatomical structures is likely to cause lung alterations including alveolar damage, interstitial edema and vascular injury responsible for the impaired gas exchange [17, 20]. It is also likely to exacerbate the inflammatory response associated with the tissue damage. The intensity of C staining was generally related to the extent of pulmonary pathological alterations with particular reference to the severity and diffusion of the inflammatory response. There was one notable exception of a patient who had massive inflammation with loss of lung architecture and yet no sign of C deposition. The reason for this observation is not apparent, but it may well be that in this case the inflammatory process is induced and sustained by pro-inflammatory cytokines known to be involved in the pulmonary inflammation in COVID-19 patients [21].

Considering that all our patients were elderly people aged more than 70, it is not surprising that they had multiple comorbidities that may have contributed to induce C activation together with SARS-CoV-2 infection. However, while this possibility cannot be excluded, it should be pointed out that the comorbidities in the majority of cases did not specifically affect the respiratory tract, and when they did in two cases, C deposits were markedly reduced compared to all the other cases. A point of concern was that the post-mortem period varying between 1 and 3 days prior to autopsies might have had an impact on the degree of tissue C deposition, but the autopsies of COVID-19 and control groups, with the only exception of case 6, were performed within the same range of post-mortem interval. An interesting finding of this study is that the C deposits were not restricted to the lungs but were also detected in kidneys and liver of the same patients, in line with the accepted view that COVID-19 is a multi-organ disease [4, 7, 19]. Both these organs shared with the lungs a common feature of predominant, though not exclusive, C localization on blood vessels suggesting the contribution of locally deposited C activation products to microvascular injury and thrombosis.

Published reports have suggested that C is mainly activated through the lectin pathway in patients infected by SARS-Cov-2 [22, 23]. The viral Spike protein has been proposed as a possible candidate to trigger the lectin pathway given the high degree of glycosylation that may enable this protein to interact with the recognition molecules of the pathway, though the evidence to support this possibility is lacking. The other viral protein that may be implicated in the activation of the lectin pathway is the N protein of SARS-Cov-2. Gao and colleagues [14] reported in a non-peer reviewed publication data indicating that N protein binds MASP2 and enhances the cleavage of C4 and C3 induced by the immobilized MBL-MASP2 complex. They also show that components of the lectin pathway are present in the pulmonary tissue of COVID-19 patients, although the protein deposits are hardly visible in the published figure, raising some doubt on the in vivo relevance of these results. Staining for MASP-2 was also observed by Magro et al [16] in the post-mortem pulmonary analysis in COVID-19 patients, but the finding was limited to one out of five patients examined and no information was given on the tissue localization of MBL and other recognition molecules of the lectin pathway. The faint or absent staining of our patients’ lung for MBL and MASP-2 cannot be attributed to the failure of the antibodies to recognize their targets as both proteins were detected in the liver cells that synthesize these proteins, but rather indicates that the lectin pathway is not the main driver of C activation in our study group. However, we cannot exclude that the lectin pathway may be involved in the promotion of thrombosis, one the hallmarks of COVID-19 disease. Eriksson and coworkers [24] has recently reported increased levels of MBL in COVID-19 patients with thrombotic episodes that were correlated with the levels of plasma D-dimers, but not with the inflammatory process and organ alterations. These data are consistent with our recent observation that MBL interacts with beta2-glycoprotein expressed on endothelial cells and activates the lectin pathway leading to MASPs-mediated cleavage of prothrombin and thrombin generation [18]. The pro-coagulant activity of MASPs may explain the beneficial effects observed in COVID-19 patients treated with Narsoplimab [25] which inhibits MASP-2, preventing C activation and thrombus formation, although the small number of treated patients and the concomitant use of other drugs represent important limitations, recognized by the authors of this study.

The positive staining of the patients’ lung for C1q points to the classical pathway as an important route of C activation likely triggered by the IgG that are widely distributed in the lung of all patients examined. Unfortunately, the immunohistochemical analysis does not permit to define the antigenic specificity of these IgG. However, it is tempting to speculate that the spike protein of SARS-CoV-2 present in several tissue samples is a potential target, although we cannot exclude that other viral and tissue antigens may also be recognized by the antibodies. The latter possibility is suggested by the reports of sequence homology between SARS-CoV-2 and human proteins-derived peptides published by different groups [26-28] and is supported in this study by the finding of Spike protein-independent localization of IgG in the lung of COVID-19 patients. An important implication of these observations is that, because of cross-reactivity of human and viral proteins, the antibody response to SARS-CoV-2 may contribute to the pathogenesis of the disease, causing tissue damage possibly as a result of C activation. A similar conclusion was reached some years ago by Yang and colleagues [29] who were able to detect antibodies to primary human pulmonary endothelial cells and human pulmonary epithelial cell line in patients with SARS-associated coronavirus infection. More recently, Wang and colleagues [30] have reported a substantial increase of autoantibodies directed against immunomodulatory molecules responsible for dysregulation of immune functions in COVID-19 patients.

The detection of IgG and C1q in kidneys and liver is consistent with similar finding in the lung specimens and supports the conclusion that the classical pathway is the prevailing route of C activation, further amplified by the involvement of the alternative pathway. As hypothesized for C activation in the lung, the complex of antibody and Spike protein documented in both kidney [31, 32] and liver [33] may be the initial trigger of C activation followed by the intervention of additional factors that contribute to potentiate C activation with the progression of the disease severity.

In conclusion, this work has shown that C deposits can be detected in post-mortem biopsy specimens of various organs including lung, kidney and liver collected from COVID-19 patients, supporting the accepted view that this is a multi-organ disease. In addition, evidence has been provided that C is preferentially activated through the classical pathway with the important contribution of the alternative pathway. Taken together, these data further support the involvement of C in COVID-19 pathogenesis and the use of therapeutic agents to prevent C-dependent tissue damage and sustained inflammatory response associated with this severe infectious disease.

## Data Availability

all data referred in the manuscript are available

## Contributors

Paolo Macor designed the study, wrote the manuscript, analyzed the tissue sections and prepared tables and figures;

Paolo Durigutto performed the immunofluorescence staining and analyzed the results, prepared figures;

Alessandro Mangogna organized the study and analyzed the tissue sections

Rossana Bussani performed autopsies and collected tissue samples;

Stefano D’Errico collaborate with Committee of the Regione Friuli Venezia Giulia for ethical approval;

Martina Zanon collected clinical and laboratory data;

Nicola Pozzi contributed to design the study and to write the manuscript;

PierLuigi Meroni assisted in the clinical evaluation of the patients and provided important suggestion for the design of the study

Francesco Tedesco designed the study, wrote the manuscript, analysed the tissue sections.

## Declaration of Interests

There are no conflicts of interest.

## Acknowledgments

Grants from the Italian Ministry of Health (COVID-2020-12371808) to PLM and National Institutes of Health HL150146 to NP are gratefully acknowledged.

## Notes

### Competing Interest Statement

The authors have declared no competing interest.

### Author Declarations

This work was approved by the Ethical Committee of the Regione Friuli Venezia Giulia, Italy (Prot. N. 0025523 / P / GEN/ ARCS).

